# Perspectives on mental illness stigma among African immigrant pregnant and post-partum women in an urban setting: a brief report

**DOI:** 10.1101/2021.11.06.21266011

**Authors:** Aderonke Bamgbose Pederson, Elizabeth Waldron, Inger Burnett-Zeigler, Crystal Clark, Lynette Lartey, Katherine Wisner

## Abstract

**Purpose:** This study assessed the perspectives of pregnant and post-partum African immigrant women on mental illness.

**Methods:** We conducted a focus group session (N=14) among pregnant and postpartum African immigrant women in June 2020. We used an inductive driven thematic analysis to identify themes related to mental health stigma.

**Results:** Five core themes emerged: conceptualization of mental health, community stigmatizing attitudes, biopsychosocial stressors, management of mental health and methods to reduce stigma.

**Conclusion:** Understanding the perspectives of pregnant African immigrant women at the intersection of their race, ethnicity, gender and migration is necessary to improve engagement with mental health services.

## Introduction

The African immigrant population increased by nearly 90% since 2000, yet they remain underrepresented in health care research (1, 2). Pregnant and postpartum African immigrant women experience their mental health in pregnancy at the intersection of their race, ethnicity, gender and migrant status. This period is a vulnerable time that leaves mothers highly susceptible to psychiatric disorders (3, 4).

In the US, 13-19% of women who give birth experience postpartum depression (3, 5). Despite the similarity in prevalence of postpartum depression among White, Black and Latina people, utilization of mental health services is lower among African immigrants (6, 7). Rates of depression treatment are particularly low among Black compared to White pregnant women (8). Moreover, migrant status affects health outcomes; for example, Canadian-born women experienced significantly different risk factors for postpartum depression compared to immigrant women (9). Data on mental health and associated stigma among pregnant and post-partum immigrant women in the US has focused more on Latina rather than Black immigrant women (10).

To address mental illness stigma and improve engagement in mental health services (11-14), dedicated studies are necessary in the peripartum and postpartum period for African immigrant women. We assessed the perspectives of pregnant and postpartum African immigrants on the conceptualization of their emotional health, views on mental health stigma and the underlying religious and cultural beliefs influencing mental health service use.

## Methods

### Ethical Consideration

The institutional review board at Northwestern University approved this study. Written and informed consent was completed.

### Setting, Participants and Study Design

#### Setting

The study was completed in partnership with the United African Organization (UAO). UAO provided social services to African immigrants. The Northwestern University team included a psychiatrist and a research coordinator.

#### Participants

The focus group and survey participants (N=14) inclusion criteria were:1) identifying as an African immigrant, 2) pregnant or postpartum within the past 12 months, 3) English speaking, and 4) 18 years of age or older. Participants were recruited using convenience sampling. We held one focus group and participants also completed a brief survey on perspectives on mental health.

#### Recruitment

Flyers and brochures were placed in the community organization offices. Community partners used text messages and word of mouth to publicize the study. Interested community members were self-referred.

#### Interview Procedure and Protocol

The semi-structured interview guide and survey components were developed by a multidisciplinary team including academic and community partners to ensure culturally acceptable language in discussing mental health. We developed the interview guide and survey based on the existing literature and knowledge gaps around perspectives on mental illness (8).

The questionnaire was completed prior to the focus group and all study activities lasted approximately 75 minutes. Due to the COVID 19 pandemic, the study activity was held online via videoconferencing. Participants received $30 compensation for their time. The focus group was facilitated by an experienced facilitator (a first generation African immigrant female), and notes were taken to aid reliability and validity. Probes and member checking were used to expand ideas and verify meaning expressed by participants (15). All participant’ information was de-identified to ensure confidentiality.

### Data Analysis

The focus group questions are presented in Table 1. We used a grounded theory thematic approach (16, 17). NVivo software, a qualitative analysis tool, was used. The focus group was audio recorded and transcribed using Rev, an authenticated platform for transcription. We employed five iterative steps (16): initial review, line coding, organization of meaning units, discussion and final review of consistency between coders. Intercoder reliability (K > 0.80) was reached. Coders applied the codebook to the full transcript.

**Table 1.** Focus group questions

| Focus Group Questions |
| --- |
| 1) What is emotional wellness? |
| 2) What is your understanding of changes in emotional wellness including stress or distress that can happen while pregnant or after pregnancy? |
| 3) What are your perceptions of your community's understanding of stigma of emotional wellness while pregnant or after pregnancy? Do you believe emotional wellness of mothers is looked down upon in our society/community? In what ways? |
| 4) What is the definition of mental health stigma? |
| 5) Tell us about personal stigma of emotional wellness while pregnant or after pregnancy: |
| What are your personal beliefs about emotional wellness during or after pregnancy, beliefs about emotional wellness treatment – including **views on taking medication** - while pregnant or breastfeeding or in general? |
| 6) Do people experience shame about mental illness at home or at work or in public settings while pregnant or after a pregnancy? What can reduce the negative attitudes or feelings of shame? |
| 7) What do you think would help reduce stigma around having emotional stress or challenges? Would you participate in these strategies such as meeting peers who have mental health challenges (direct contact), education about mental health using presentations or videos, entertainment or theatre, mental health awareness campaigns? What are some barriers to participating? |

### Assessments

We developed a *socio-demographic questionnaire* to assess age group, education, marital status, citizenship status, income, employment status and insurance status. The *Patient Health Questionnaire (PHQ2)* is a validated two-item depression screening measure (18). This measure has a high positive predictive value (18). We also developed a *brief questionnaire* to assess specific beliefs about mental illness such as beliefs that mental illness is caused by sin or a moral failure as well as views on medications (19). The main analysis in this study was conducted using a focus group interview guide, see table 1.

## Results

Among the African immigrant women (n=14) in the study, 50.0% of respondents reported being pregnant (n=7; 5 in the first trimester) and three (21.4%) participants had a positive screen on the PHQ-2. Participants’ characteristics are shown in table 2.

**Table 2.**
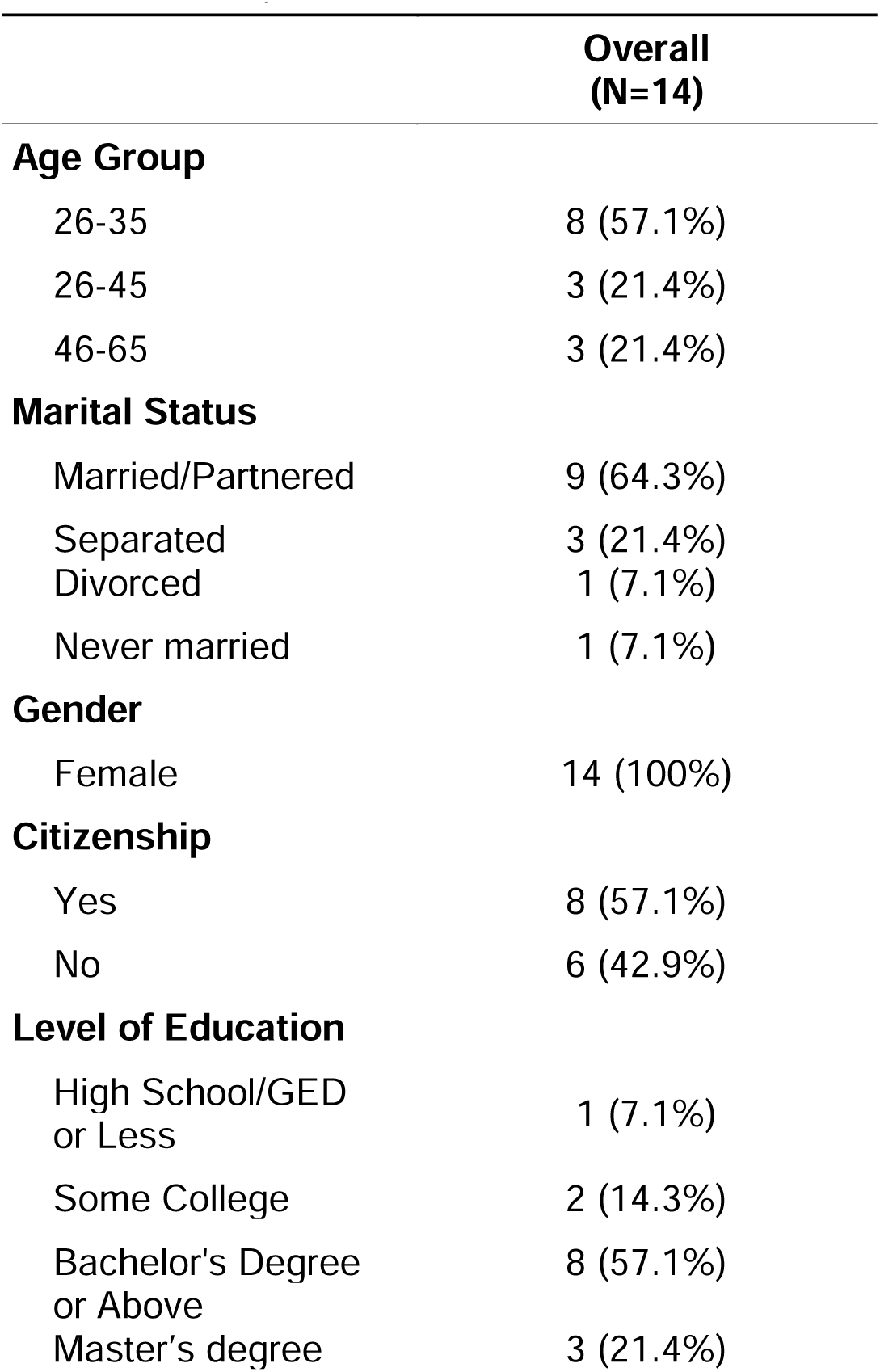
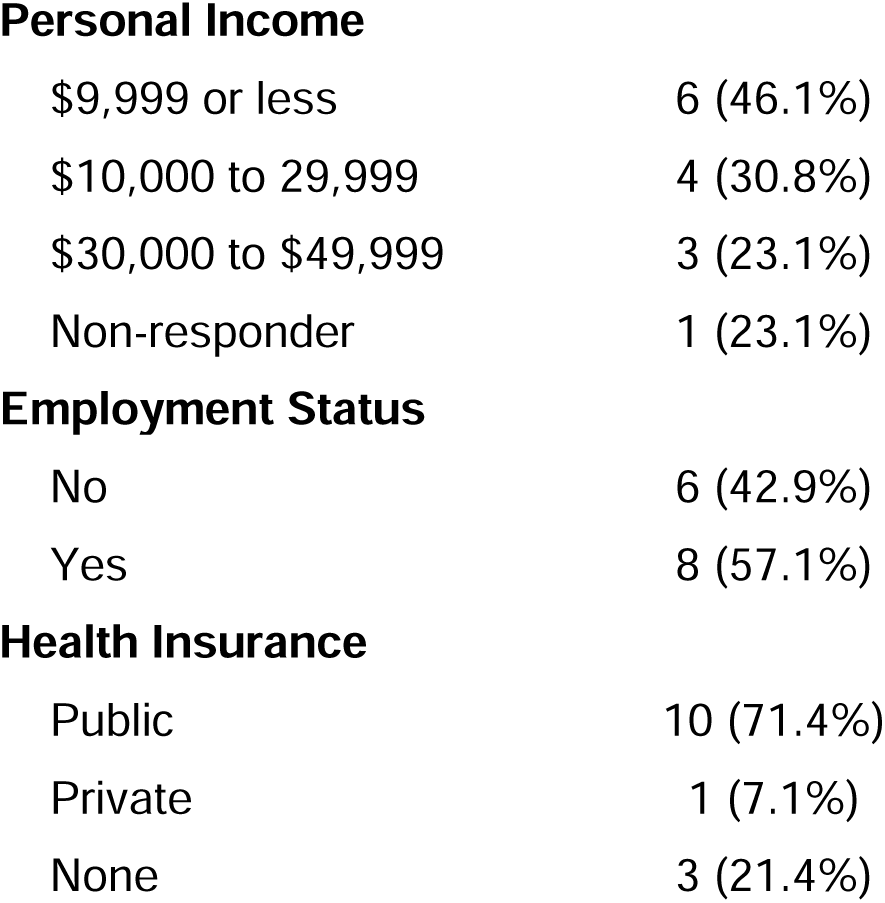
Characteristics of sample Participant characteristics

|  | **Overall**<br>**(N=14)** |
| --- | --- |
| **Age Group** |  |
| 26-35 | 8 (57.1%) |
| 26-45 | 3 (21.4%) |
| 46-65 | 3 (21.4%) |
| **Marital Status** |  |
| Married/Partnered | 9 (64.3%) |
| Separated | 3 (21.4%) |
| Divorced | 1 (7.1%) |
| Never married | 1 (7.1%) |
| **Gender** |  |
| Female | 14 (100%) |
| **Citizenship** |  |
| Yes | 8 (57.1%) |
| No | 6 (42.9%) |
| **Level of Education** |  |
| High School/GED or Less | 1 (7.1%) |
| Some College | 2 (14.3%) |
| Bachelor's Degree or Above | 8 (57.1%) |
| Master's degree | 3 (21.4%) |
| **Personal Income** |  |
| \$9,999 or less | 6 (46.1%) |
| \$10,000 to 29,999 | 4 (30.8%) |
| \$30,000 to \$49,999 | 3 (23.1%) |
| Non-responder | 1 (23.1%) |
| **Employment Status** |  |
| No | 6 (42.9%) |
| Yes | 8 (57.1%) |
| **Health Insurance** |  |
| Public | 10 (71.4%) |
| Private | 1 (7.1%) |
| None | 3 (21.4%) |

**Table 3.**
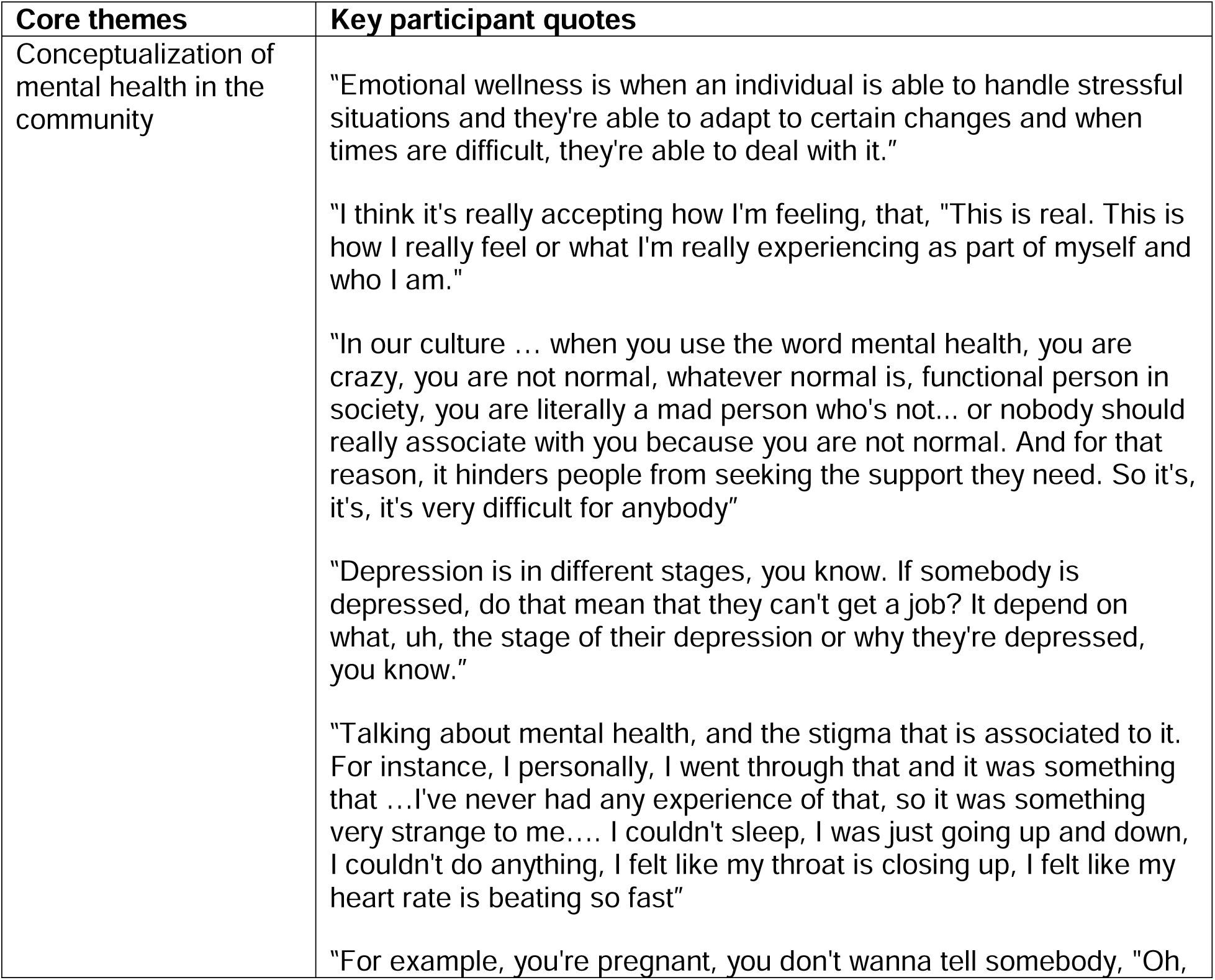
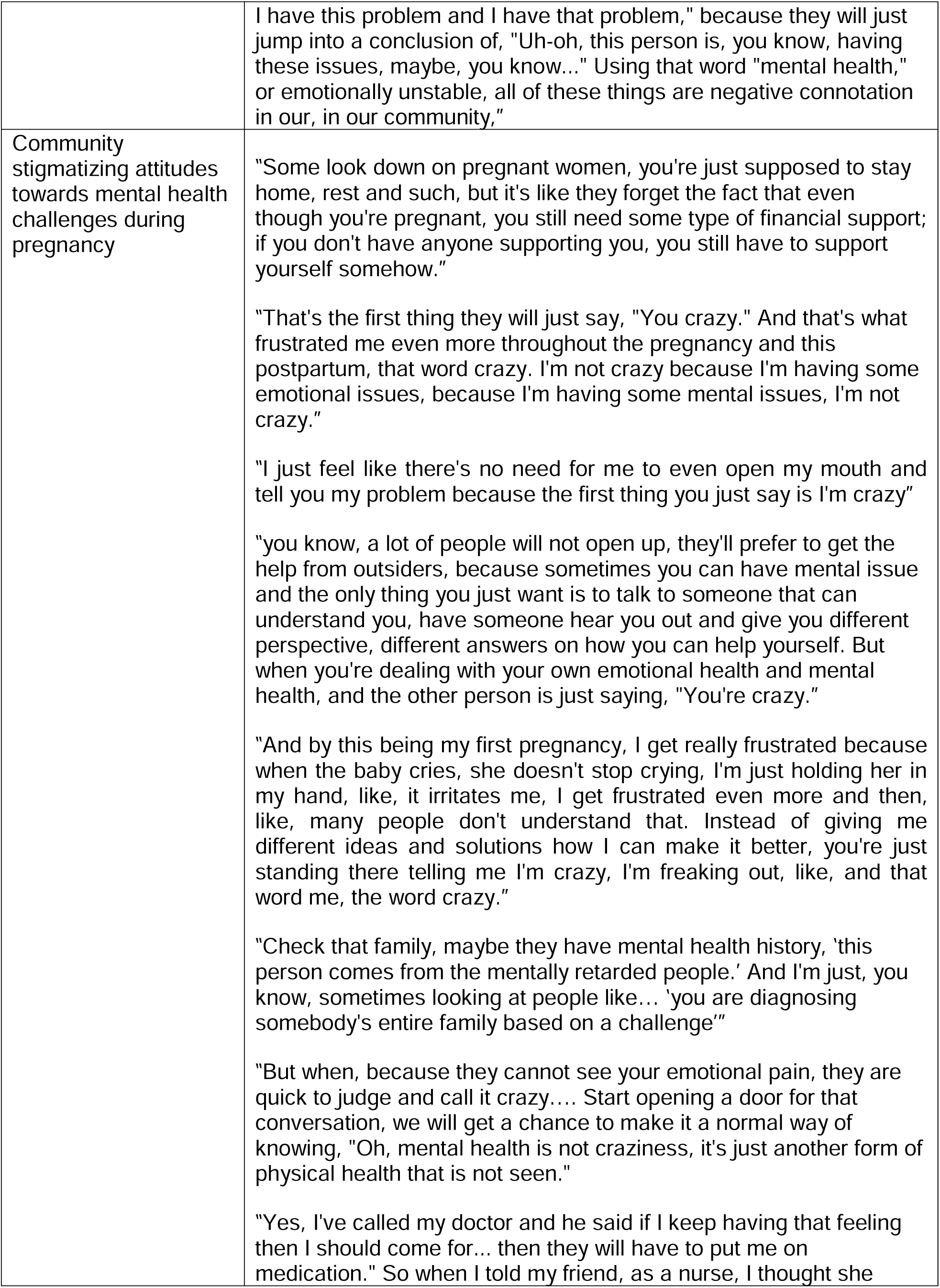
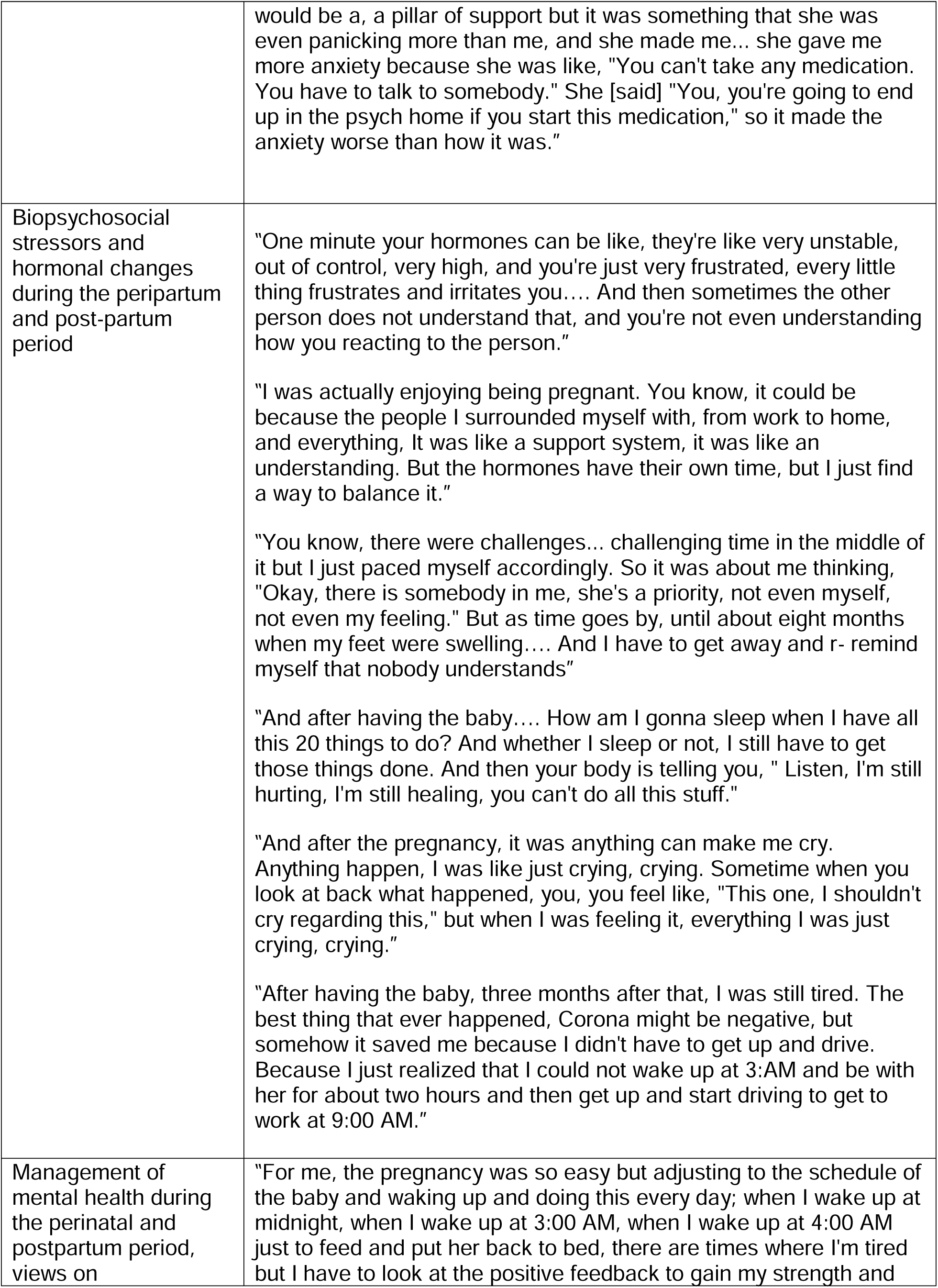
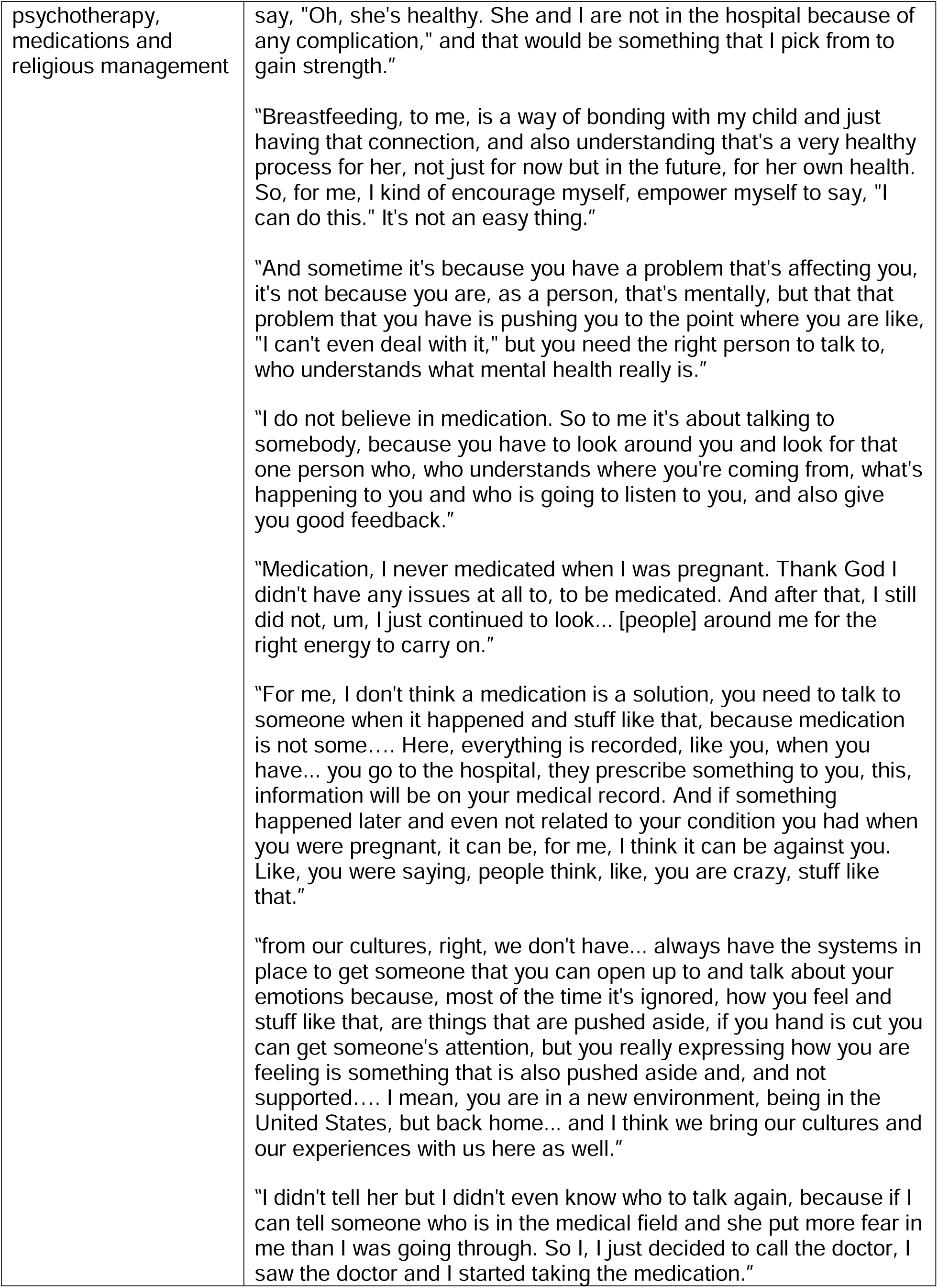
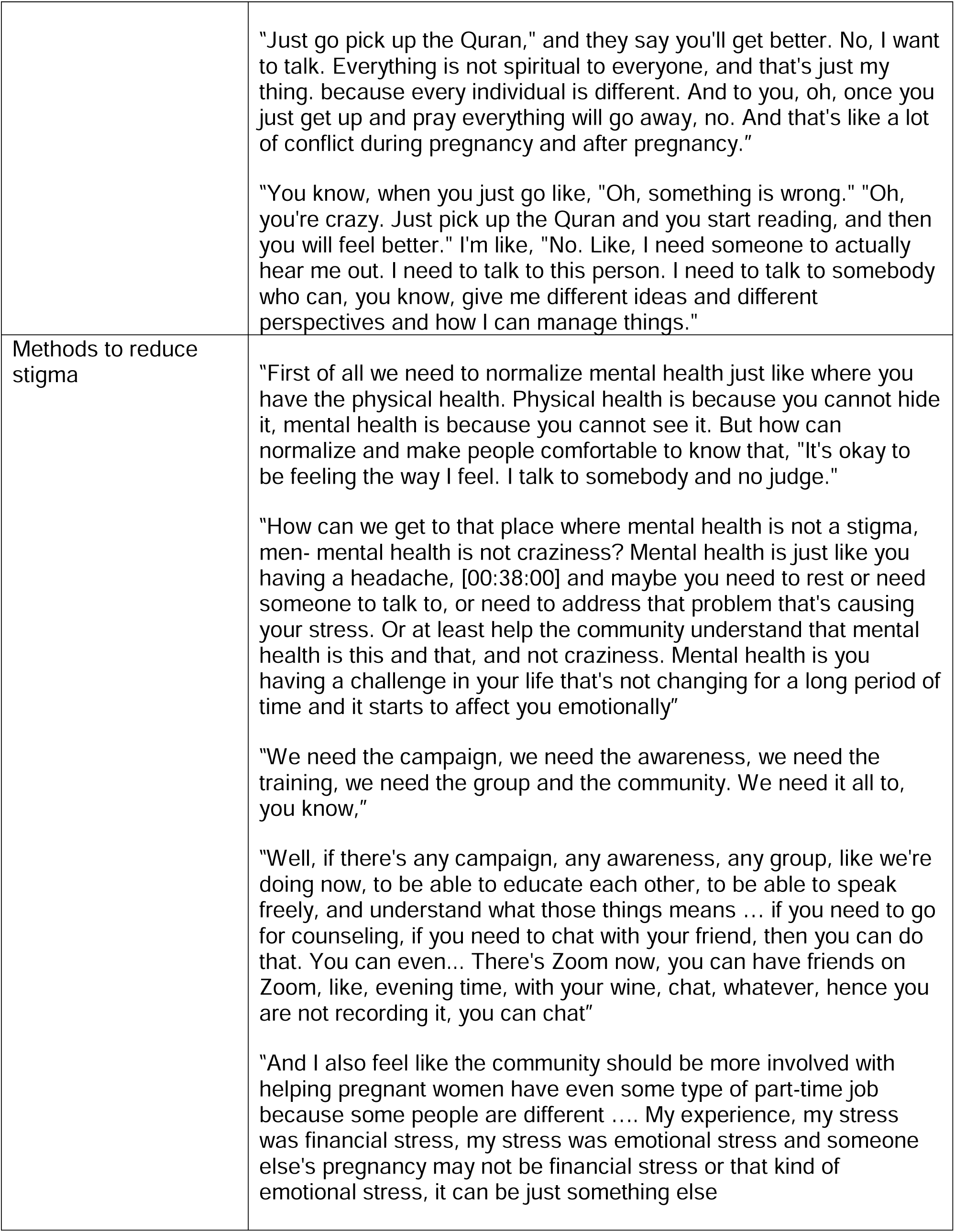
Core themes and representative quotes

| Core themes | Key participant quotes |
| --- | --- |
| Conceptualization of mental health in the community | "Emotional wellness is when an individual is able to handle stressful situations and they're able to adapt to certain changes and when times are difficult, they're able to deal with it."<br>"I think it's really accepting how I'm feeling, that, "This is real. This is how I really feel or what I'm really experiencing as part of myself and who I am."<br>"In our culture ... when you use the word mental health, you are crazy, you are not normal, whatever normal is, functional person in society, you are literally a mad person who's not... or nobody should really associate with you because you are not normal. And for that reason, it hinders people from seeking the support they need. So it's, it's, it's very difficult for anybody"<br>"Depression is in different stages, you know. If somebody is depressed, do that mean that they can't get a job? It depend on what, uh, the stage of their depression or why they're depressed, you know."<br>"Talking about mental health, and the stigma that is associated to it. For instance, I personally, I went through that and it was something that ...I've never had any experience of that, so it was something very strange to me.... I couldn't sleep, I was just going up and down, I couldn't do anything, I felt like my throat is closing up, I felt like my heart rate is beating so fast"<br>"For example, you're pregnant, you don't wanna tell somebody, "Oh, |
|  | I have this problem and I have that problem," because they will just jump into a conclusion of, "Uh-oh, this person is, you know, having these issues, maybe, you know..." Using that word "mental health," or emotionally unstable, all of these things are negative connotation in our, in our community," |
| Community stigmatizing attitudes towards mental health challenges during pregnancy | "Some look down on pregnant women, you're just supposed to stay home, rest and such, but it's like they forget the fact that even though you're pregnant, you still need some type of financial support; if you don't have anyone supporting you, you still have to support yourself somehow."<br>"That's the first thing they will just say, "You crazy." And that's what frustrated me even more throughout the pregnancy and this postpartum, that word crazy. I'm not crazy because I'm having some emotional issues, because I'm having some mental issues, I'm not crazy."<br>"I just feel like there's no need for me to even open my mouth and tell you my problem because the first thing you just say is I'm crazy"<br>"you know, a lot of people will not open up, they'll prefer to get the help from outsiders, because sometimes you can have mental issue and the only thing you just want is to talk to someone that can understand you, have someone hear you out and give you different perspective, different answers on how you can help yourself. But when you're dealing with your own emotional health and mental health, and the other person is just saying, "You're crazy."<br>"And by this being my first pregnancy, I get really frustrated because when the baby cries, she doesn't stop crying, I'm just holding her in my hand, like, it irritates me, I get frustrated even more and then, like, many people don't understand that. Instead of giving me different ideas and solutions how I can make it better, you're just standing there telling me I'm crazy, I'm freaking out, like, and that word me, the word crazy."<br>"Check that family, maybe they have mental health history, 'this person comes from the mentally retarded people.' And I'm just, you know, sometimes looking at people like... 'you are diagnosing somebody's entire family based on a challenge'"<br>"But when, because they cannot see your emotional pain, they are quick to judge and call it crazy.... Start opening a door for that conversation, we will get a chance to make it a normal way of knowing, "Oh, mental health is not craziness, it's just another form of physical health that is not seen."<br>"Yes, I've called my doctor and he said if I keep having that feeling then I should come for... then they will have to put me on medication." So when I told my friend, as a nurse, I thought she |
|  | would be a, a pillar of support but it was something that she was even panicking more than me, and she made me... she gave me more anxiety because she was like, "You can't take any medication. You have to talk to somebody." She [said] "You, you're going to end up in the psych home if you start this medication," so it made the anxiety worse than how it was." |
| Biopsychosocial stressors and hormonal changes during the peripartum and post-partum period | "One minute your hormones can be like, they're like very unstable, out of control, very high, and you're just very frustrated, every little thing frustrates and irritates you.... And then sometimes the other person does not understand that, and you're not even understanding how you reacting to the person."<br>"I was actually enjoying being pregnant. You know, it could be because the people I surrounded myself with, from work to home, and everything, It was like a support system, it was like an understanding. But the hormones have their own time, but I just find a way to balance it."<br>"You know, there were challenges... challenging time in the middle of it but I just paced myself accordingly. So it was about me thinking, "Okay, there is somebody in me, she's a priority, not even myself, not even my feeling." But as time goes by, until about eight months when my feet were swelling.... And I have to get away and r- remind myself that nobody understands"<br>"And after having the baby.... How am I gonna sleep when I have all this 20 things to do? And whether I sleep or not, I still have to get those things done. And then your body is telling you, " Listen, I'm still hurting, I'm still healing, you can't do all this stuff."<br>"And after the pregnancy, it was anything can make me cry. Anything happen, I was like just crying, crying. Sometime when you look at back what happened, you, you feel like, "This one, I shouldn't cry regarding this," but when I was feeling it, everything I was just crying, crying."<br>"After having the baby, three months after that, I was still tired. The best thing that ever happened, Corona might be negative, but somehow it saved me because I didn't have to get up and drive. Because I just realized that I could not wake up at 3:AM and be with her for about two hours and then get up and start driving to get to work at 9:00 AM." |
| Management of mental health during the perinatal and postpartum period, views on | "For me, the pregnancy was so easy but adjusting to the schedule of the baby and waking up and doing this every day; when I wake up at midnight, when I wake up at 3:00 AM, when I wake up at 4:00 AM just to feed and put her back to bed, there are times where I'm tired but I have to look at the positive feedback to gain my strength and |
| psychotherapy, medications and religious management | say, "Oh, she's healthy. She and I are not in the hospital because of any complication," and that would be something that I pick from to gain strength."<br>"Breastfeeding, to me, is a way of bonding with my child and just having that connection, and also understanding that's a very healthy process for her, not just for now but in the future, for her own health. So, for me, I kind of encourage myself, empower myself to say, "I can do this." It's not an easy thing."<br>"And sometime it's because you have a problem that's affecting you, it's not because you are, as a person, that's mentally, but that that problem that you have is pushing you to the point where you are like, "I can't even deal with it," but you need the right person to talk to, who understands what mental health really is."<br>"I do not believe in medication. So to me it's about talking to somebody, because you have to look around you and look for that one person who, who understands where you're coming from, what's happening to you and who is going to listen to you, and also give you good feedback."<br>"Medication, I never medicated when I was pregnant. Thank God I didn't have any issues at all to, to be medicated. And after that, I still did not, um, I just continued to look... [people] around me for the right energy to carry on."<br>"For me, I don't think a medication is a solution, you need to talk to someone when it happened and stuff like that, because medication is not some.... Here, everything is recorded, like you, when you have... you go to the hospital, they prescribe something to you, this, information will be on your medical record. And if something happened later and even not related to your condition you had when you were pregnant, it can be, for me, I think it can be against you. Like, you were saying, people think, like, you are crazy, stuff like that."<br>"from our cultures, right, we don't have... always have the systems in place to get someone that you can open up to and talk about your emotions because, most of the time it's ignored, how you feel and stuff like that, are things that are pushed aside, if you hand is cut you can get someone's attention, but you really expressing how you are feeling is something that is also pushed aside and, and not supported.... I mean, you are in a new environment, being in the United States, but back home... and I think we bring our cultures and our experiences with us here as well."<br>"I didn't tell her but I didn't even know who to talk again, because if I can tell someone who is in the medical field and she put more fear in me than I was going through. So I, I just decided to call the doctor, I saw the doctor and I started taking the medication." |
|  | "Just go pick up the Quran," and they say you'll get better. No, I want to talk. Everything is not spiritual to everyone, and that's just my thing. because every individual is different. And to you, oh, once you just get up and pray everything will go away, no. And that's like a lot of conflict during pregnancy and after pregnancy."<br>"You know, when you just go like, "Oh, something is wrong." "Oh, you're crazy. Just pick up the Quran and you start reading, and then you will feel better." I'm like, "No. Like, I need someone to actually hear me out. I need to talk to this person. I need to talk to somebody who can, you know, give me different ideas and different perspectives and how I can manage things." |
| Methods to reduce stigma | "First of all we need to normalize mental health just like where you have the physical health. Physical health is because you cannot hide it, mental health is because you cannot see it. But how can normalize and make people comfortable to know that, "It's okay to be feeling the way I feel. I talk to somebody and no judge."<br>"How can we get to that place where mental health is not a stigma, men- mental health is not craziness? Mental health is just like you having a headache, [00:38:00] and maybe you need to rest or need someone to talk to, or need to address that problem that's causing your stress. Or at least help the community understand that mental health is this and that, and not craziness. Mental health is you having a challenge in your life that's not changing for a long period of time and it starts to affect you emotionally"<br>"We need the campaign, we need the awareness, we need the training, we need the group and the community. We need it all to, you know,"<br>"Well, if there's any campaign, any awareness, any group, like we're doing now, to be able to educate each other, to be able to speak freely, and understand what those things means ... if you need to go for counseling, if you need to chat with your friend, then you can do that. You can even... There's Zoom now, you can have friends on Zoom, like, evening time, with your wine, chat, whatever, hence you are not recording it, you can chat"<br>"And I also feel like the community should be more involved with helping pregnant women have even some type of part-time job because some people are different .... My experience, my stress was financial stress, my stress was emotional stress and someone else's pregnancy may not be financial stress or that kind of emotional stress, it can be just something else" |

### Summary of focus group and brief questionnaire results

Five core themes related to mental health and mental health stigma emerged:

Theme 1. *Conceptualization of mental health in the community*. Some respondents described mental health as an emotional experience, possessing resilience and having good functional wellbeing. The word “mental” was seen as having negative connotations. Some respondents (n = 4, 28.57%) believe depression is not a medical illness, and some respondents (n = 3, 21.4%) reported people with depression are dangerous, while others (n =7, 50%) reported people with depression are unpredictable.

Theme 2. *Community stigmatizing attitudes towards mental health challenges during pregnancy*. Several respondents shared their experience of judgment. People with emotional struggles were seen as “crazy” or “mentally retarded.” There was majority consensus that pregnant women should not take medication while pregnant (n = 10, 71.4%). Some respondents reported feelings of sadness or depression are a moral failure (7 [50%]), a result of sin (6 [42.9%]) or caused by evil spirits (5 [35.6%]).

Theme 3. *Biopsychosocial stressors and hormonal changes during the peripartum and post-partum period*. There was general consensus that pregnancy was connected to changes in the emotional state of a person. Stressors included biological and psychosocial aspects of stress – hormones, lack of sleep, the overwhelming nature of personal and professional responsibilities.

Theme 4. *Management of mental health during the perinatal and postpartum period*. Respondents offered several ways to manage mental health in the context of pregnancy including self-motivation, religious resources and talking to others (both in the community and professionally). While 50.0% of respondents reported that they would call 911 or seek immediate help if someone was having suicidal thoughts, five (35.7%) said they would never or rarely seek immediate help.

Theme 5. *Methods to reduce stigma*. One respondent described mental health as invisible and difficult to “normalize.” Respondents suggested public health campaigns and awareness groups in the community, similar to the focus group forum. In particular, one respondent described the need to support pregnant women due to the added psychosocial stressors that come with pregnancy.

## Discussion

Our main finding was that stigma towards mental health was associated with the label of being “crazy” or being judged. In addition, medication was not viewed as acceptable, but there was openness to activities such as support from the community and psychotherapy. There was acceptance of the biopsychosocial aspects of pregnancy (including hormonal changes); despite this acknowledgement, most respondents had stigmatizing views of medications. Majority of respondents (64.3%) reported they would rarely or never take prescription medications. And some respondents endorsed beliefs that sadness or depression is a moral failure, a sin or caused by evil spirits. Past studies show that stigmatizing beliefs similar to these led to low utilization of mental health services and worse mental health outcomes related to higher morbidity and mortality (20, 21).

Previous studies have also shown that pregnant African immigrant women had better self-rated physical and mental health compared to US-born and Caribbean-born pregnant Black women (10), likely due to the healthy migrant effect (6, 22). The healthy migrant effect refers to the concept that migrants tend to have better health status than people of similar backgrounds in the host country (23); however, over time, likely due to the effect of racism, gender and migration, this apparent advantage is diminished (23, 24). In our sample, the willingness to seek professional mental health services, especially pharmacotherapy was particularly low.

There are higher somatization rates among African immigrants, compared to US born Black people; African immigrants tend to focus on physical health symptoms and prefer primary care health professionals (6, 7). In our study, respondents endorsed anxiety and low mood symptoms, such as difficulty sleeping, elevated heart rate, tearfulness, irritability and generally feeling overwhelmed. Due to somatization, pregnant African immigrant women are at elevated risk of health care professionals not recognizing their symptoms. Therefore, they may experience further delay in initiation and engagement in mental health services in pregnancy, where early intervention is critical (4, 25).

Our finding should be considered in the context of the study design; though the sample size was small, there were several areas of consensus around conceptualization of mental illness and associated stigma.

## Conclusion

This study shows conceptualization of mental illness, stigmatizing views and specific negative attitudes towards medication use among peripartum and postpartum African immigrant women. Understanding the perspectives of pregnant African immigrant women at the intersection of their race, ethnicity, gender and migrant status is necessary to address mental illness stigma that leads to low utilization of mental health services within our health systems (6, 7).

## Data Availability

All data produced in the present work are contained in the manuscript

